# High proportions of post-exertional malaise and orthostatic intolerance in people living with post-COVID-19 condition: the PRIME post-COVID study

**DOI:** 10.1101/2023.08.17.23294204

**Authors:** Demi M. E. Pagen, Maarten Van Herck, Céline J. A. van Bilsen, Stephanie Brinkhues, Kevin Konings, Casper D. J. den Heijer, Martijn A. Spruit, Christian J. P. A. Hoebe, Nicole H. T. M. Dukers-Muijrers

## Abstract

**Background:** Exercise-based treatments can be harmful in people who were SARS-CoV-2 positive and living with post-COVID-19 condition (PL-PCC) and who have post-exertional malaise (PEM) or orthostatic intolerance (OI). Nevertheless, PEM and OI are not routinely assessed by clinicians. We estimated PEM and OI proportions in PL-PCC, as well in people not living with PCC (PnL-PCC) and negatives (i.e., never reported a SARS-CoV-2 positive test), and identified associated factors.

**Methods:** Participants from the PRIME post-COVID study were included. PEM and OI were assessed using validated questionnaires. PCC was defined as feeling unrecovered after SARS-CoV-2 infection. Multivariable regression analyses to study PEM and OI were stratified for sex.

**Results:** Data from 3,783 participants was analyzed. In PL-PCC, proportion of PEM was 48.1% and 41.2%, and proportion of OI was 29.3% and 27.9% in women and in men, respectively. Proportions were higher in PL-PCC compared to negatives, for PEM in women OR=4.38 [95%CI:3.01-6.38]; in men OR=4.78 [95%CI:3.13-7.29]; for OI in women 3.06 [95%CI:1.97-4.76]; in men 2.71 [95%CI:1.75-4.21]. Associated factors were age ≤60 years, ≥1 comorbidities and living alone.

**Conclusions:** High proportions of PEM and OI are observed in PL-PCC. Standard screening for PEM and OI is recommended in PL-PCC, to promote appropriate therapies.

Trial registration ClinicalTrials.gov identifier:NCT05128695

## Background

There is an urgent need for information on optimal care and treatment options for people living with post-COVID-19 condition (PL-PCC) (1). PL-PCC are suffering from substantial, persistent symptoms after a SARS-CoV-2 infection. Experienced symptoms are very heterogenous, including fatigue, dyspnea and cognitive disfunctions, among others (2, 3). A link between post-COVID-19 condition (PCC) and myalgic encephalomyelitis or chronic fatigue syndrome (ME/CFS) has been made (4, 5), particularly based on the similarities in the presence or relapsing of unexplained symptoms, like disabling fatigue, exhaustion, difficulty thinking, pain, exercise intolerance, and other symptoms (6, 7).

Post-exertional malaise (PEM) has previously been described in PCC (3, 8), and is a cardinal feature of ME/CFS (9, 10). PEM subscribes the abnormal worsening of various symptoms (which can be fatigue) and loss of energy following minimal physical or cognitive stressors, or other triggers that would have been tolerated normally before disease onset (6). PEM has been found more prevalent in women than in men (11) and infections can initiate PEM (12). Another comorbid with ME/CFS is the postural orthostatic tachycardia syndrome (POTS), which generally causes orthostatic intolerance (13). The majority of the POTS patients are women as well (14).

Rehabilitation of PL-PCC is often focused on applying exercise-based protocols, especially early on in the COVID-19 pandemic, as early reports of cases were derived from deconditioned hospitalized cases (8, 15). However, the relation between physical activity and PCC is not well understood; with some studies describing improved symptoms and others symptom exacerbation (16). Also, some PCC patients are offered cognitive behavioral therapy (CBT), but curative merit is criticized in line with concerns in ME/CFS patients (17). The presence of PEM or OI in people with PCC has important implications for their treatment options, as people can be intolerant to exercise, cognitive stressors, or to upright position. There is evidence that exercise-based protocols can be harmful (18). To date, the prevalence of PEM and OI in PL-PCC is not well known (19), but likely substantial.

This observational cohort study, called the Prevalence, Risk factors, and Impact Evaluation post-COVID study (PRIME post-COVID), estimated the proportion of PEM and OI in PL-PCC and people who were SARS-CoV-2 positive and not living with PCC (PnL-PCC) and adults who never reported a positive SARS-CoV-2 test (further referred to as negatives). Furthermore, we identified relevant subgroups that are more prone to have PEM or OI, and described the occurrence of fatigue or other symptoms that may accompany PEM or OI in PL-PCC.

## Methods

### Study design

The design and recruitment of the PRIME post-COVID study has been published previously (20). In brief, an observational open cohort study was set up with assessments on various health-conditions and health-factors. Invitees were adults tested for COVID-19 with a valid test result and email address, recorded in the public health registry in Southern Limburg, the Netherlands. The longitudinal character enabled additional data collection moments. After completing the baseline questionnaire (December 2021), participants were invited to participate in a follow-up questionnaire (August 2022).

### Participants

In total, 12,453 initial participants were invited to complete the follow-up questionnaire. Data were collected using the online MWM2 application of market research platform Crowdtech (ISO 27001 certified). Participants who likely represented another person than the intended invitee (reported inconsistent information regarding sex and test result compared to the baseline questionnaire) were excluded.

### Data collection

The follow-up questionnaire covered demographics (to construct variables on age, sex, level of education, and urbanity of living area), date and result of last COVID-19 test, physical health (height and weight to construct body mass index (BMI), and comorbidity), and smoking behavior.

Additionally, the questionnaire included the validated DePaul Symptom Questionnaire Post-Exertional Malaise (DSQ-PEM) (21) and four items from the DePaul Symptom Questionnaire-2 (DSQ-2) regarding OI (22, 23), and experienced symptoms (44 pre-listed) with severity scores (range 1-10). Based on the reported symptoms, participants were categorized into:

- Did not experience any symptoms now
- Experienced fatigue only
- Experienced fatigue and at least one other symptom
- Experienced multiple symptoms except fatigue

Frequencies and proportions of these categories were reported in PL-PCC with and without PEM or OI. The questionnaire further included a question whether people felt recovered or not felt recovered since their first recorded SARS-CoV-2 infection.

### Classification of COVID-19 test result

For people invited at baseline, COVID-19 test result was known in the national test registry. Additionally, participants self-reported SARS-CoV-2 infections in both questionnaires. Participants were classified as SARS-CoV-2 negatives when no positive test result was reported in both baseline (registry and self-report) and follow-up questionnaire (self-report). However, we have to acknowledge that at the time of the follow-up questionnaire (August 2022), the chance that people were truly negative and never had been infected before is small. Nevertheless, to retain readability and clarity of this paper, people who never reported a positive SARS-CoV-2 test will be referred to as negatives.

Participants who reported at least one positive test result (i.e., in baseline or follow-up) were classified as SARS-CoV-2 positive.

### Outcome variables

#### Post-exertional malaise

In the DSQ-PEM, respondents rated five items over the previous six months on frequency (never, sometimes, about half the time, most of the time, always) and severity (no, mild, moderate, severe, very severe) on a 5-point Likert scale. The five items were “A dead, heavy feeling after starting to exercise”, “Next day soreness or fatigue after non-strenuous, everyday activities”, “Mentally tired after the slightest effort”, “Minimum exercise makes you physically tired”, and “Physically drained or sick after mild activity”. A score on frequency of about half of the time to always and a score on severity of moderate to very severe on the same item on any of the five items is indicative for PEM (21). Additionally, in people who had PEM, a sum score (range 4-40; minimum of 4 due to the threshold for having PEM) of frequency (range 0-4) and severity (0–4) of the five items was calculated as severity measure (24).

#### Orthostatic intolerance

OI was measured using four items selected from the DSQ-2. Respondents rated the four items over the previous six months on frequency (never, sometimes, about half the time, most of the time, always) and severity (no, mild, moderate, severe, very severe) on a 5-point Likert scale. The four items were “Rapid heartbeat after standing”, “Blurred or tunnel vision after standing”, “Gray or blacking after standing”, and “Inability to tolerate an upright position”. A score on frequency of about half of the time to always and a score on severity of moderate to very severe on the same item on any of the four items is indicative for OI. These four items were selected as these are used in the various classifications of ME/CFS to define OI (22, 23). Also for OI, in people who had OI, a severity sum score (range 4-32; minimum of 4 due to the threshold for having OI) was calculated based on frequency (range 0-4) and severity (range 0-4) of the four items.

Moreover, co-occurrence of PEM and OI was described by reporting proportions of participants having: no PEM nor OI; only OI; only PEM; both PEM and OI.

### Post-COVID condition definition and study population in current analyses

Several PCC definitions have previously been studied within the PRIME post-COVID study (25). In the current study, we aimed to inform clinicians on the proportions of PEM and OI in the PCC population as well as in the general population. As we sought to inform clinical practice, we considered it appropriate to use the PCC definition of not feeling recovered, as this most likely reflects the population who would present to medical care. Not feeling recovered has also been used as PCC definition in various previous studies (25–31).

As sensitivity analyses, we presented various other PCC definitions and estimated PEM and OI proportions (25). These other PCC definitions included:

1. Having ≥1 of all 44 pre-listed symptom
2. Having ≥1 symptoms that were significantly more often reported in positives than in negatives (in data of baseline questionnaire)
3. Having ≥1 of the selected symptoms in definition 2 AND with a severity score of ≥5 points (cut-off of 5 was used according to the mean of scores; range 1-10).

Besides, sensitivity analyses were performed by presenting PEM and OI proportions stratified for months since first reported positive SARS-CoV-2 test (3-6, 6-9, 9-12, 12-18 and longer than 18 months ago) in PL-PCC using the PCC definition of not feeling recovered.

### Associated factors

Several demographics (sex, age, level of education, living alone), physical (obesity, comorbidities), lifestyle (current or former smoking behavior), and environmental (urbanity of living area) factors have been selected as factors possibly associated with PEM and OI. These subgroup characteristics are often known to the treating physician and might be of use to indicate high prevalence subgroups. Age was dichotomized into 18-60 and 60+ age groups, based on the age distribution of our study population. Level of education was categorized into people being practically (i.e., no, lower general, lower vocational, general secondary, and secondary vocational education) or theoretically (i.e., higher general, pre-university, higher professional, and scientific education) trained. Being obese was defined as having a BMI≥30 kg/m^2^. Urbanity of living area was based on postal code and categorized into: (very) strongly urban, moderately urban, little urban, rural.

### Statistical analysis

Participants who reported ME/CFS or fibromyalgia before their SARS-CoV-2 infection were excluded from the analyses to limit a possible risk of overestimating proportions of PEM and OI. People who tested SARS-CoV-2 positive less than three months before questionnaire completion were also excluded, because of the PCC definition window. Other studies found that women more often had PEM and OI than men (11, 14). As this was confirmed in our study population for PEM, subsequent analyses were stratified by sex.

Proportions and 95% confidence intervals (CI) were calculated for PEM and OI in PL-PCC, PnL-PCC, and negatives. Associations with age, smoking behavior, living alone, urbanity of living area, having obesity, or comorbidities were performed using multivariable logistic regression analyses. We also tested for effect modification between these factors and PCC-group. In these regression analyses, PnL-PCC were excluded. Independent-samples Mann-Whitney U test was used to test whether PEM and OI severity scores differed between PL-PCC and negatives. Analyses were performed using Statistical Package for Social Sciences (SPSS; version 27.0, IBM, Armonk, USA). A p-value of <0.05 was considered statistically significant.

### Ethical statement and trial registry

The PRIME-post COVID study was waived by the Medical Ethical Committee of Maastricht University Medical Centre+ (METC2021-2884). This study was registered at Clinical Trials.gov Protocol Registration and Results System (NCT05128695).

## Results

Of the invitees (n=12,453), 4,201 (60.4%) had complete data. Of the people who tested SARS-CoV-2 positive, 253 were excluded as they reported ME/CFS or fibromyalgia before SARS-CoV-2 infection or were tested less than three months before questionnaire completion. The population in analyses consisted of n=955 PL-PCC, n=2,174 PnL-PCC, and n=654 negatives (Figure 1).

**Figure 1.**
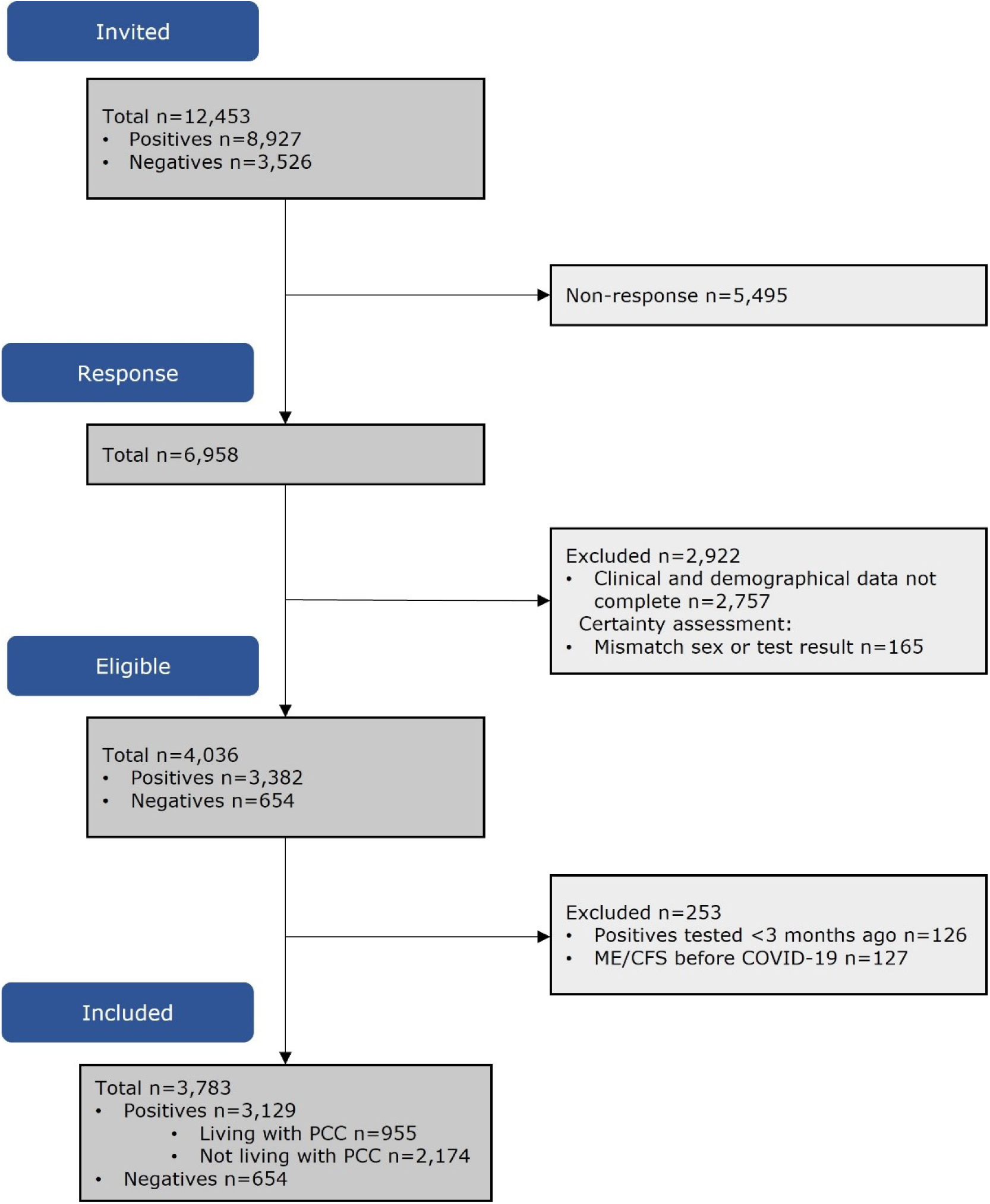
Flow chart of participants included in analyses

Groups differed regarding sex, age, educational level, BMI, comorbidities and living alone (Table 1).

**Table 1.**
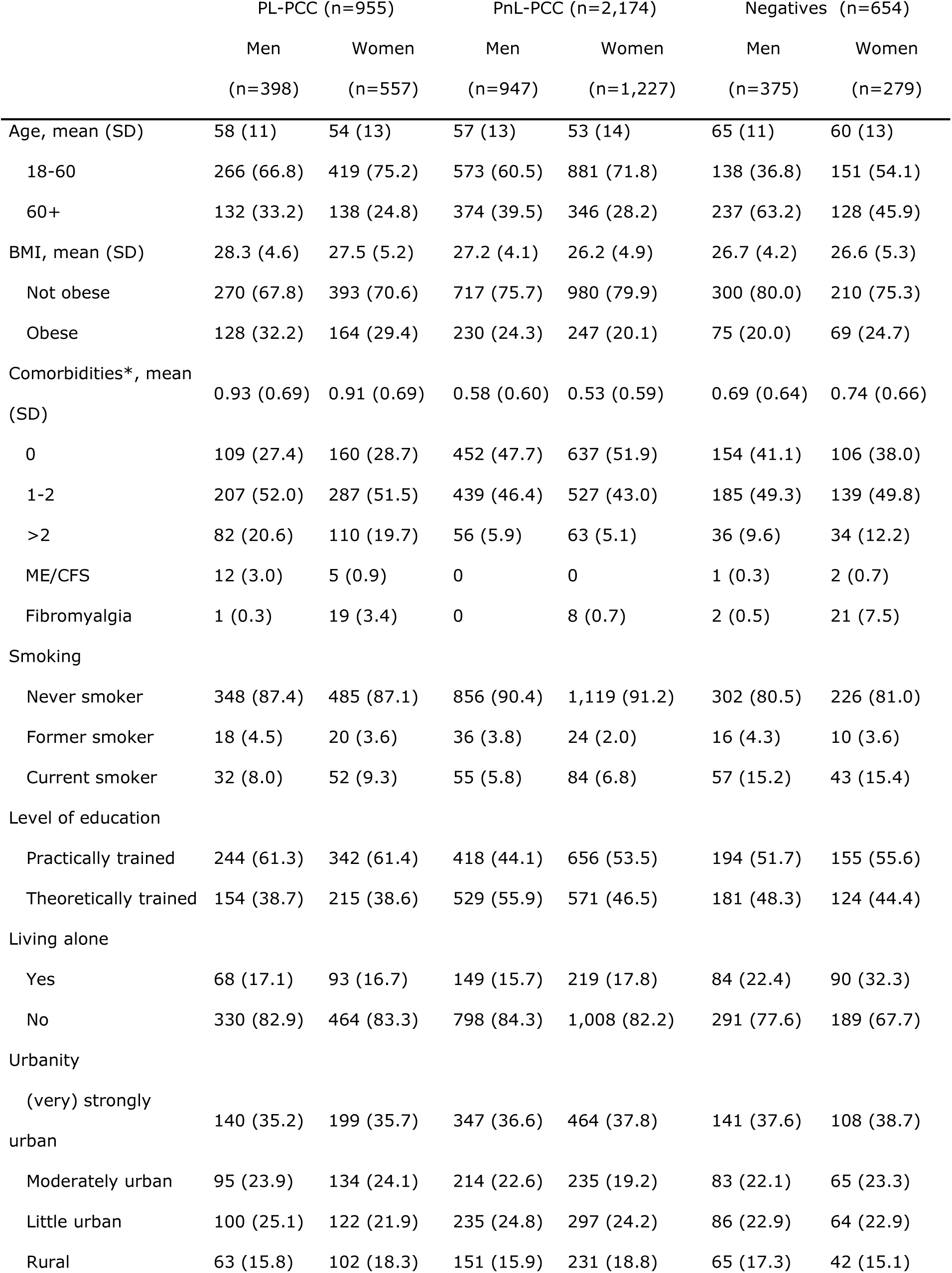
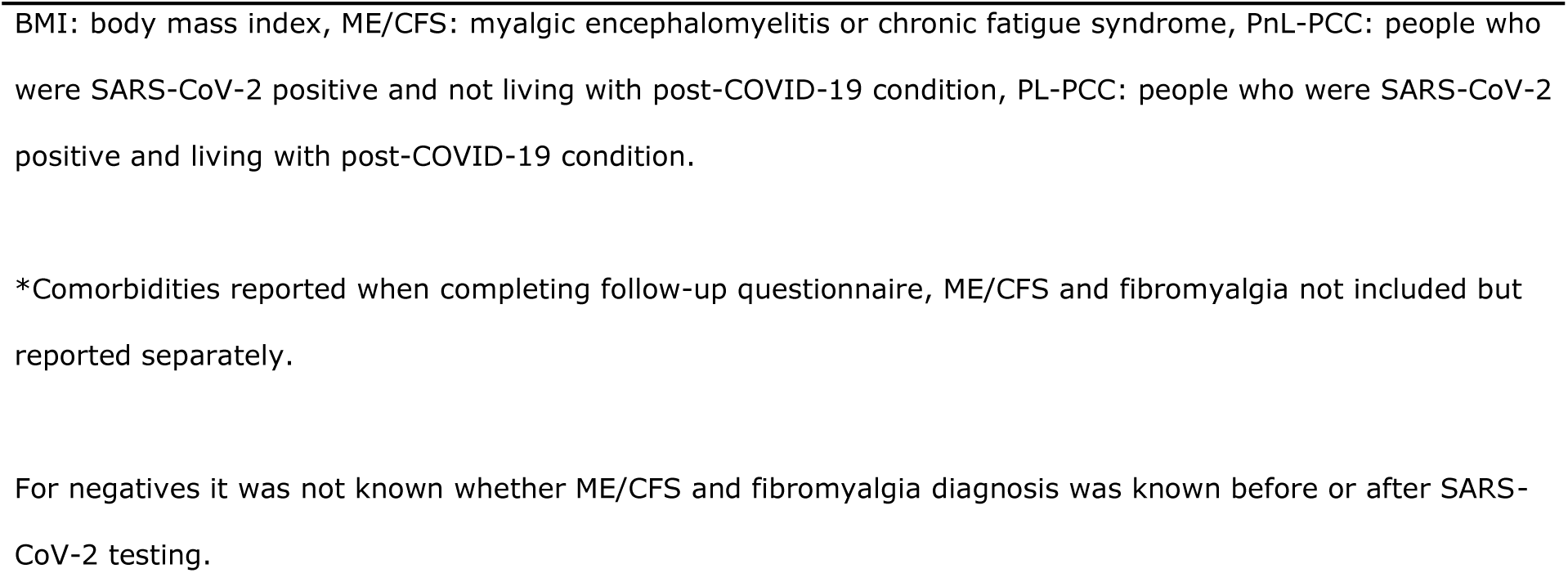
Population characteristics, stratified for people who were SARS-CoV-2 positive and are living with post-COVID-19 condition (PL-PCC), people who were SARS-CoV-2 positive and not living with post-COVID-19 condition (PnL-PCC) and negatives.

|  | PL-PCC (n=955) |  | PnL-PCC (n=2,174) |  | Negatives (n=654) |  |
| --- | --- | --- | --- | --- | --- | --- |
|  | Men | Women | Men | Women | Men | Women |
|  | (n=398) | (n=557) | (n=947) | (n=1,227) | (n=375) | (n=279) |
| Age, mean (SD) | 58 (11) | 54 (13) | 57 (13) | 53 (14) | 65 (11) | 60 (13) |
| 18-60 | 266 (66.8) | 419 (75.2) | 573 (60.5) | 881 (71.8) | 138 (36.8) | 151 (54.1) |
| 60+ | 132 (33.2) | 138 (24.8) | 374 (39.5) | 346 (28.2) | 237 (63.2) | 128 (45.9) |
| BMI, mean (SD) | 28.3 (4.6) | 27.5 (5.2) | 27.2 (4.1) | 26.2 (4.9) | 26.7 (4.2) | 26.6 (5.3) |
| Not obese | 270 (67.8) | 393 (70.6) | 717 (75.7) | 980 (79.9) | 300 (80.0) | 210 (75.3) |
| Obese | 128 (32.2) | 164 (29.4) | 230 (24.3) | 247 (20.1) | 75 (20.0) | 69 (24.7) |
| Comorbidities*, mean (SD) | 0.93 (0.69) | 0.91 (0.69) | 0.58 (0.60) | 0.53 (0.59) | 0.69 (0.64) | 0.74 (0.66) |
| 0 | 109 (27.4) | 160 (28.7) | 452 (47.7) | 637 (51.9) | 154 (41.1) | 106 (38.0) |
| 1-2 | 207 (52.0) | 287 (51.5) | 439 (46.4) | 527 (43.0) | 185 (49.3) | 139 (49.8) |
| >2 | 82 (20.6) | 110 (19.7) | 56 (5.9) | 63 (5.1) | 36 (9.6) | 34 (12.2) |
| ME/CFS | 12 (3.0) | 5 (0.9) | 0 | 0 | 1 (0.3) | 2 (0.7) |
| Fibromyalgia | 1 (0.3) | 19 (3.4) | 0 | 8 (0.7) | 2 (0.5) | 21 (7.5) |
| Smoking |  |  |  |  |  |  |
| Never smoker | 348 (87.4) | 485 (87.1) | 856 (90.4) | 1,119 (91.2) | 302 (80.5) | 226 (81.0) |
| Former smoker | 18 (4.5) | 20 (3.6) | 36 (3.8) | 24 (2.0) | 16 (4.3) | 10 (3.6) |
| Current smoker | 32 (8.0) | 52 (9.3) | 55 (5.8) | 84 (6.8) | 57 (15.2) | 43 (15.4) |
| Level of education |  |  |  |  |  |  |
| Practically trained | 244 (61.3) | 342 (61.4) | 418 (44.1) | 656 (53.5) | 194 (51.7) | 155 (55.6) |
| Theoretically trained | 154 (38.7) | 215 (38.6) | 529 (55.9) | 571 (46.5) | 181 (48.3) | 124 (44.4) |
| Living alone |  |  |  |  |  |  |
| Yes | 68 (17.1) | 93 (16.7) | 149 (15.7) | 219 (17.8) | 84 (22.4) | 90 (32.3) |
| No | 330 (82.9) | 464 (83.3) | 798 (84.3) | 1,008 (82.2) | 291 (77.6) | 189 (67.7) |
| Urbanity |  |  |  |  |  |  |
| (very) strongly urban | 140 (35.2) | 199 (35.7) | 347 (36.6) | 464 (37.8) | 141 (37.6) | 108 (38.7) |
| Moderately urban | 95 (23.9) | 134 (24.1) | 214 (22.6) | 235 (19.2) | 83 (22.1) | 65 (23.3) |
| Little urban | 100 (25.1) | 122 (21.9) | 235 (24.8) | 297 (24.2) | 86 (22.9) | 64 (22.9) |
| Rural | 63 (15.8) | 102 (18.3) | 151 (15.9) | 231 (18.8) | 65 (17.3) | 42 (15.1) |

BMI: body mass index, ME/CFS: myalgic encephalomyelitis or chronic fatigue syndrome, PnL-PCC: people who were SARS-CoV-2 positive and not living with post-COVID-19 condition, PL-PCC: people who were SARS-CoV-2 positive and living with post-COVID-19 condition.
\*Comorbidities reported when completing follow-up questionnaire, ME/CFS and fibromyalgia not included but reported separately.
For negatives it was not known whether ME/CFS and fibromyalgia diagnosis was known before or after SARS-CoV-2 testing.

### Proportion estimates of PEM

The proportion of PEM in all positives was 23.2% (95% CI:21.2%-25.2%) in women and 17.8% (95% CI:15.8%-19.8%) in men. The proportion of PEM in PL-PCC women was 48.1% (95% CI:44.0%-52.2%). In PL-PCC men, this was lower with 41.2.0% (95% CI:36.4%-46.0%) (p=0.035) (Figure 2A). The proportion in negatives was 20.4% (95% CI:15.7%-25.1%) in women and 10.7% (95% CI:7.6%-13.8%) in men (p<0.001), and in PnL-PCC was 10.2% (95% CI:8.5%-11.9%) in women and 7.9% (95% CI:6.2%-9.6%) in men (p=0.066). The proportion of PEM in PL-PCC was between 38.6%-47.8% for women and 31.8%-41.4% for men when using other PCC definitions (Supplementary Figure 1). Proportion of PEM in PL-PCC ranged from 39.1%-56.8% in women and from 38.9%-52.5% in men by the various periods since testing (Supplementary Table 1).

**Figure 2.**
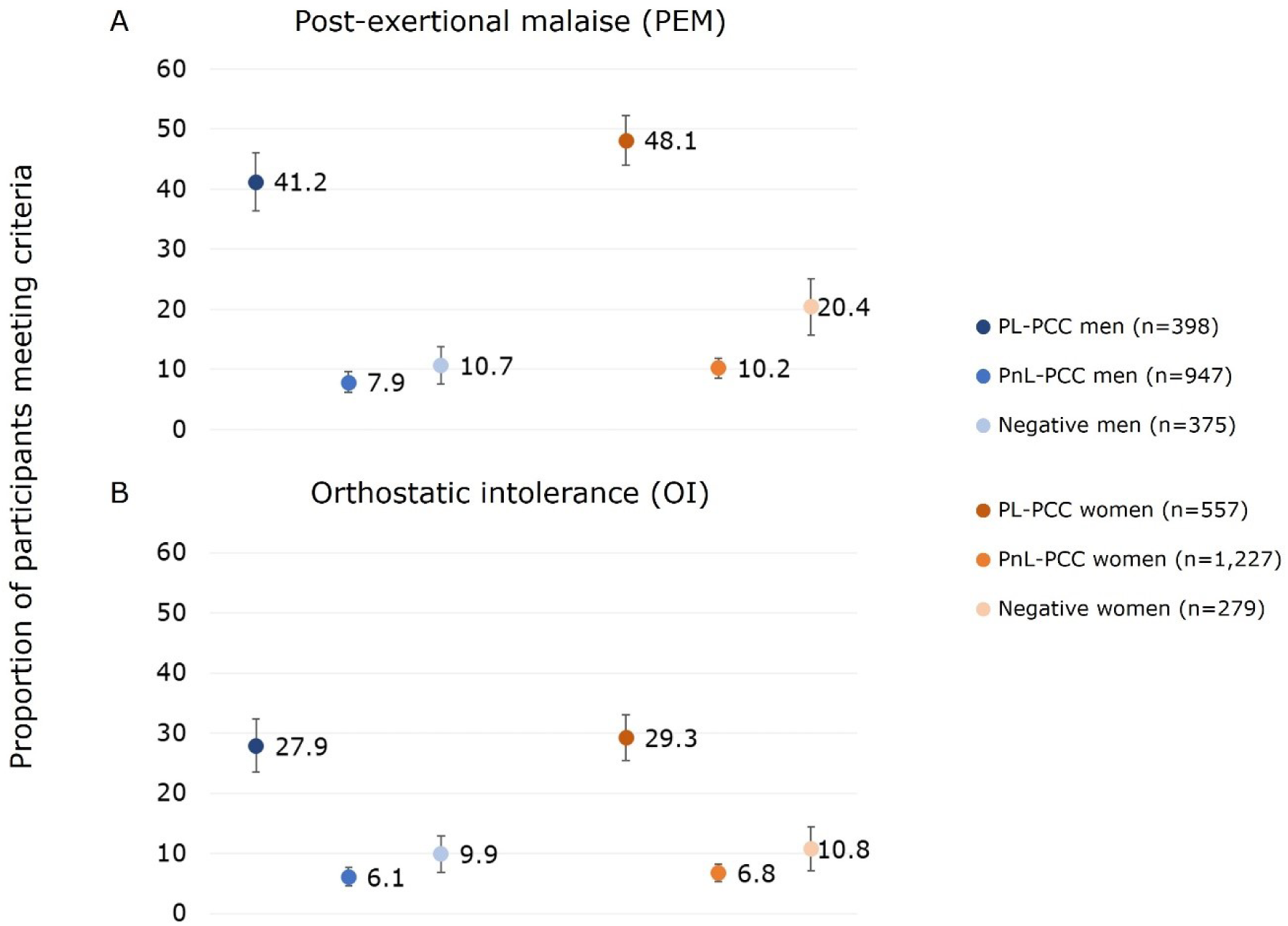
Proportion of post-exertional malaise (PEM) and orthostatic intolerance (OI) for men and women separate, in people who were SARS-CoV-2 positive and not living with post-COVID-19 condition (PnL-PCC), people who were SARS-CoV-2 positive and living with post-COVID-19 condition (PL-PCC), and negatives.

PL-PCC women had 4.38 (95% CI:3.01-6.38) higher odds of having PEM, compared to negative women, after adjusting for age, level of education, smoking behavior, living alone, urbanity of living area, obesity, and comorbidities. For PL-PCC men, the adjusted odds ratio (OR) was 4.78 (95% CI:3.13-7.29).

### Proportion estimates of OI

The proportion of OI in all positives was 13.8% (95% CI:12.2%-15.4%) in women and 12.6% (95% CI:10.8%-14.4%) in men. Proportion of OI was 29.3% (95% CI:25.5%-33.1%) in PL-PCC women and 27.9% in PL-PCC men (95% CI:23.5%-32.3%) (p=0.638) (Figure 2B). The proportion in negatives was 10.8% (95% CI:7.2%-14.4%) in women and 9.9% (95% CI:6.9%-12.9%) in men (p=0.711), and in PnL-PCC was 6.8% (95% CI:5.4%-8.2%) in women and 6.1% (95% CI:4.6%-7.6%) in men (p=0.509). The proportion of OI in PL-PCC was between 22.4%-29.1% for women and 20.4%-27.9% for men when using other PCC definitions (Supplementary Figure 1). Proportion of OI in PL-PCC ranged from 19.6%-35.1% in women and from 21.4%-32.5% in men by the various periods since testing (Supplementary Table 1).

PL-PCC women had 3.06 (95% CI:1.97-4.76) higher odds of having OI, compared to negative women, after adjusting. For PL-PCC men, the adjusted OR was 2.71 (95% CI:1.75-4.21).

### Co-occurrence between PEM and OI

PEM and OI were co-occurrent in 19.6% of the PL-PCC men and in 23.7% of the PL-PCC women. For PnL-PCC, PEM and OI were co-occurrent in 2.7% for men and 2.9% for women. In negatives, this was 5.1% in men and 7.2% in women (Figure 3).

**Figure 3.**
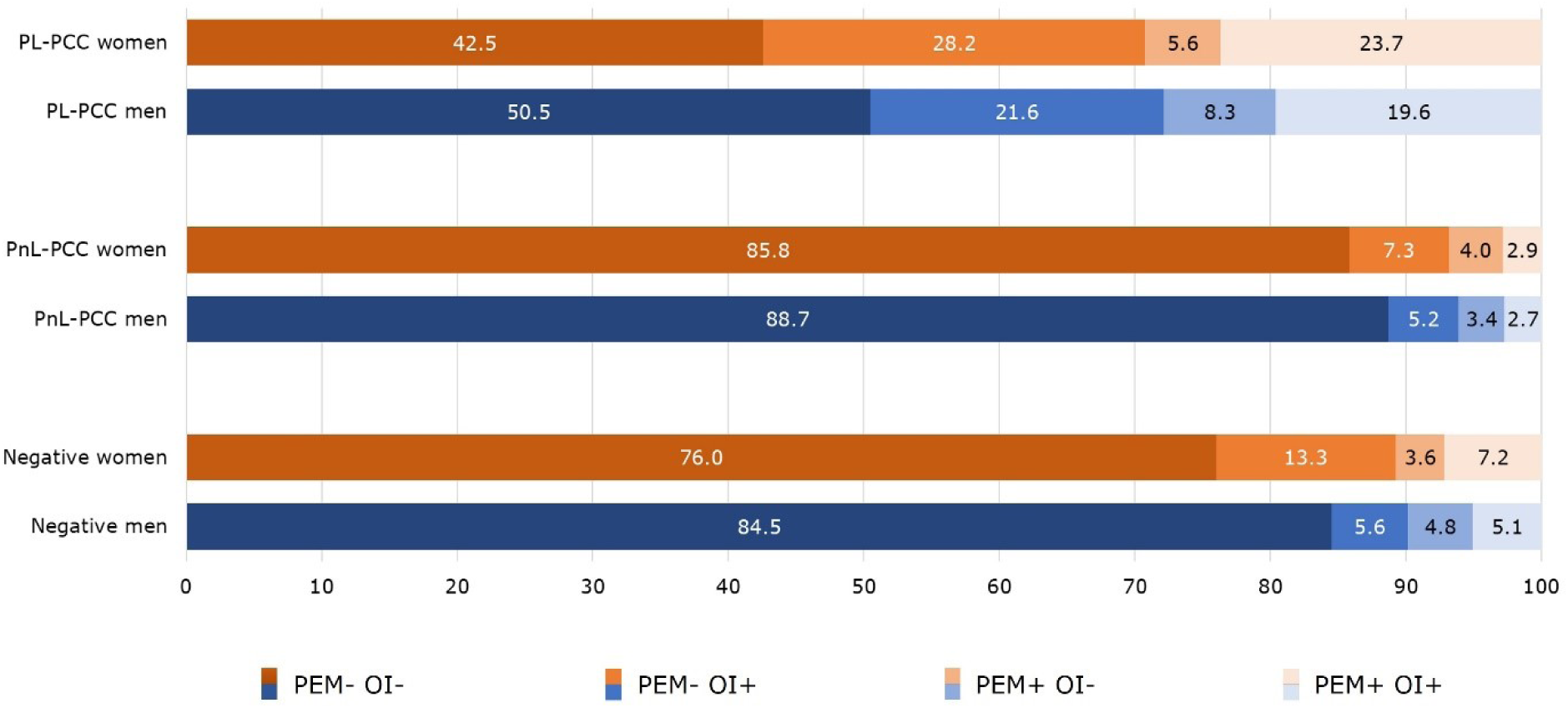
Co-occurrence of post-exertional malaise (PEM) and orthostatic intolerance (OI) in people who were SARS-CoV-2 positive and living with post-COVID-19 condition (PL-PCC), people who were SARS-CoV-2 positive and not living with post-COVID-19 condition (PnL-PCC) and negatives.

### Severity score of PEM and OI

In people who had PEM, the median PEM severity score (range 4-39) in PL-PCC was 17 in women and 18 in men. In PnL-PCC, the median PEM score was 14 in both men and women. In negatives, the median PEM score was 15 in women and 16 in men. The median PEM score was higher in PL-PCC women compared to negative women (p=0.003) (Figure 4A).

**Figure 4.**
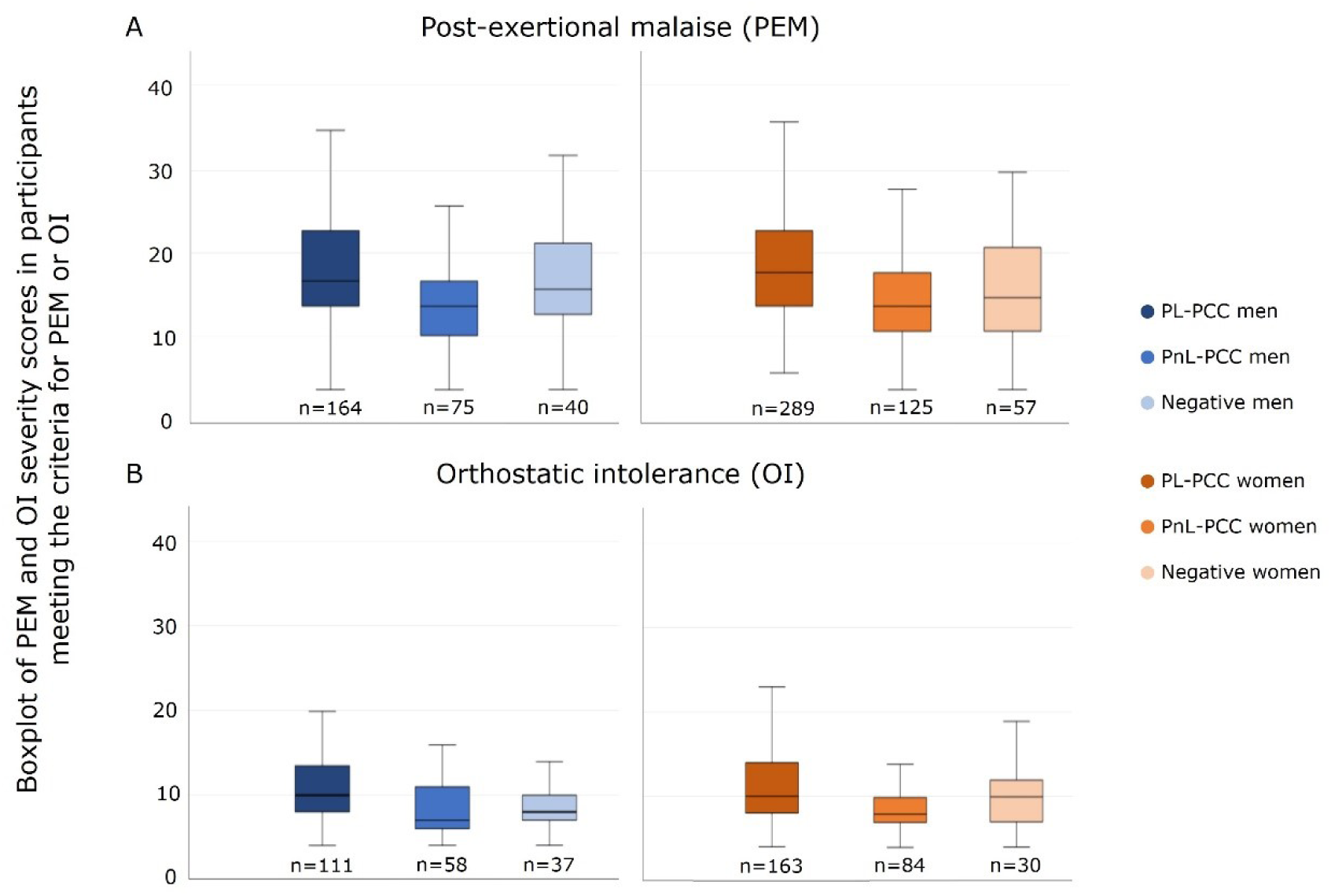
Severity score of post-exertional malaise (PEM) and orthostatic intolerance (OI) in people who had PEM or OI

In people who had OI, the median OI severity score (range 4-27) in PL-PCC was 10 in both men and women. In PnL-PCC, the median OI score was 7 in men and 8 in women. In negatives, the median OI score was 10 in women and 8 in men. The median OI severity score was higher in PL-PCC men compared to negative men (p=0.003) (Figure 4B).

### Associated factors for PEM and OI

In both men and women, factors associated with a higher risk for PEM were being aged 60 years or younger, living alone, and having at least one comorbidities; and smoking (women only) (Table 2).

**Table 2.**
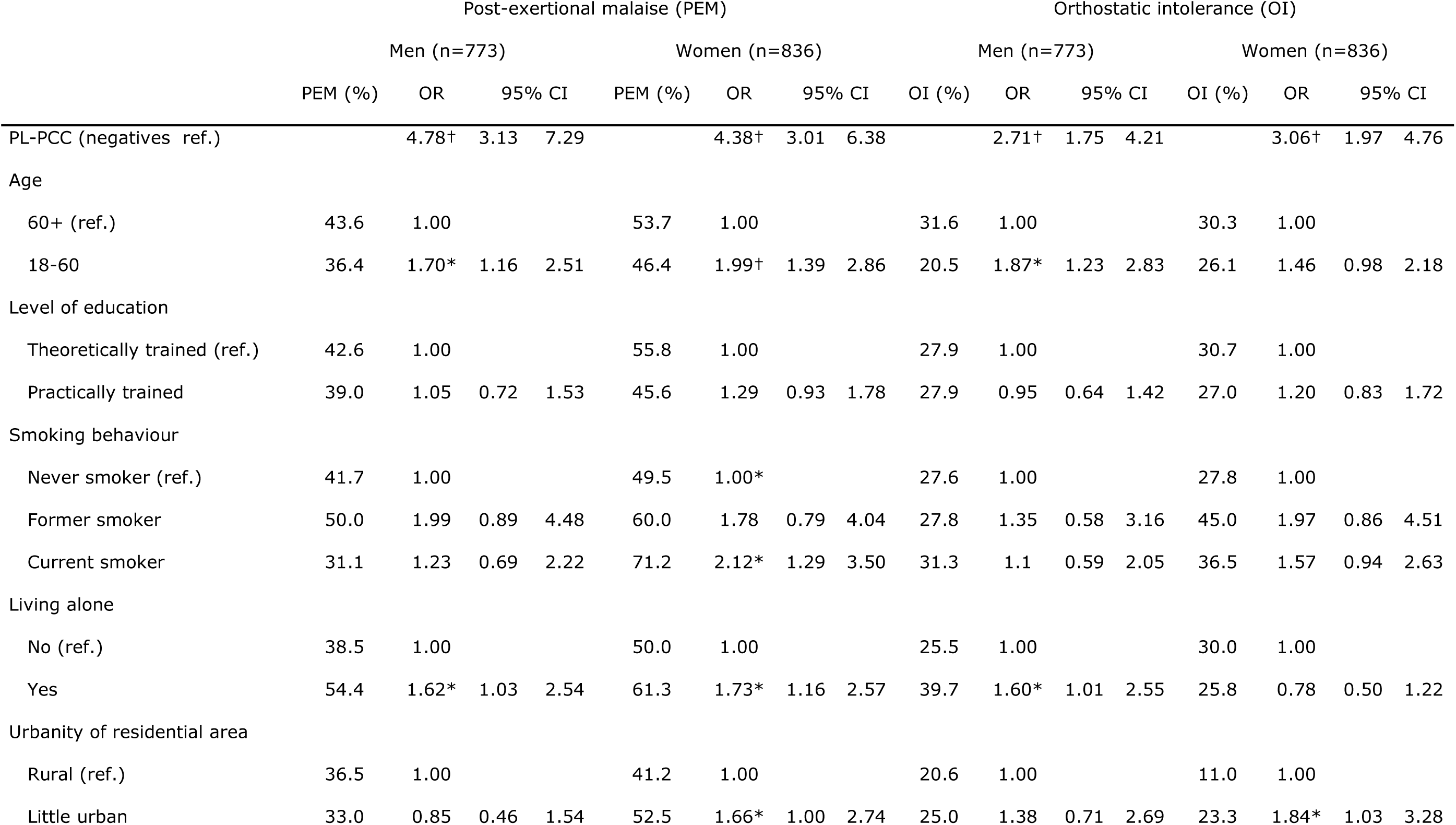
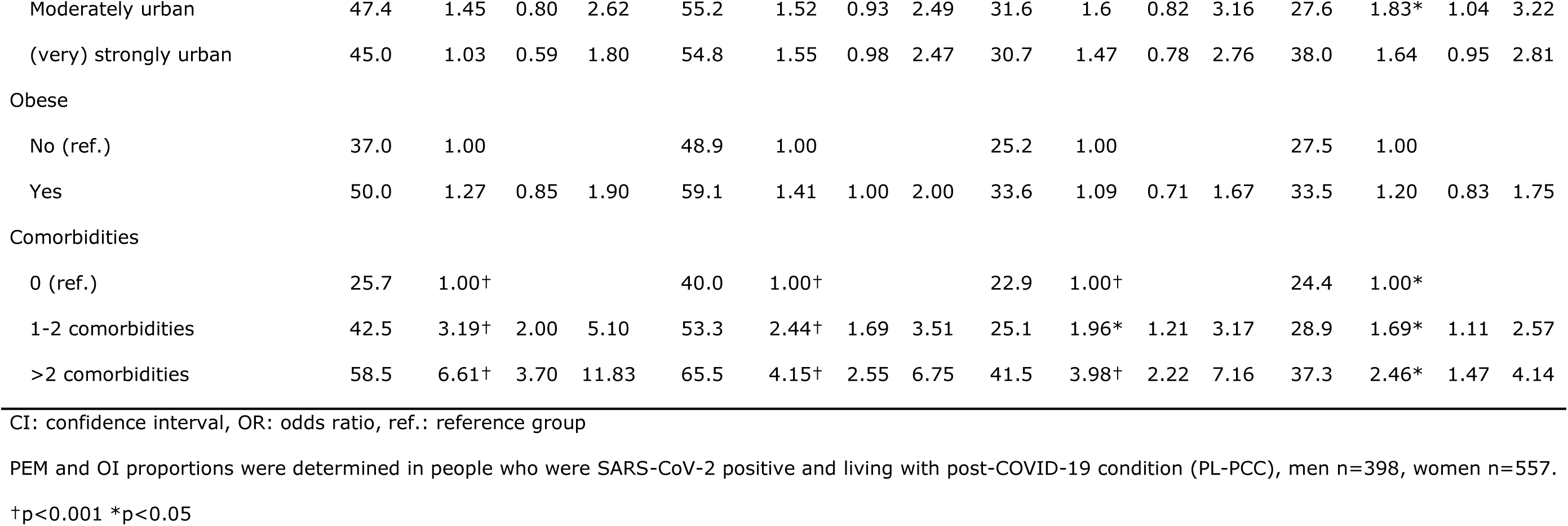
Factors associated with post-exertional malaise (PEM) or orthostatic intolerance (OI) in people who were SARS-CoV-2 positive and living with post-COVID-19 condition (PL-PCC) and negatives in multivariable regression analyses.

In both men and women, factors associated with a higher risk for OI were having at least one comorbidities, while being aged 60 years or younger and living alone was an associated factor in men only (Table 2).

Effect modification was observed between PCC group (PL-PCC versus negatives) and comorbidities, namely for both PEM and OI, the adjusted OR for comorbidities in PL-PCC was lower than in negatives (though statistically significant in both groups). Effect modification was observed between PCC group and smoking for PEM, where smoking was associated in negative men (OR_former smoker_=5.04 [95%CI:1.62-15.65]; OR_current smoker_=2.36 95%CI:1.06-5.23]), but not in PL-PCC men (OR_former smoker_=1.40 [95%CI:0.54-3.61]; OR_current smoker_=0.64 95%CI:0.29-1.38]).

### Experiencing fatigue in PL-PCC

Of the PL-PCC women who had PEM, 81.3% currently experienced fatigue (of which the majority also had other symptoms) (Figure 5A). In PL-PCC men who had PEM, 76.8% experienced fatigue (the majority also had other symptoms); thus about one in five PL-PCC with PEM did not report fatigue (Figure 5B).

**Figure 5.**
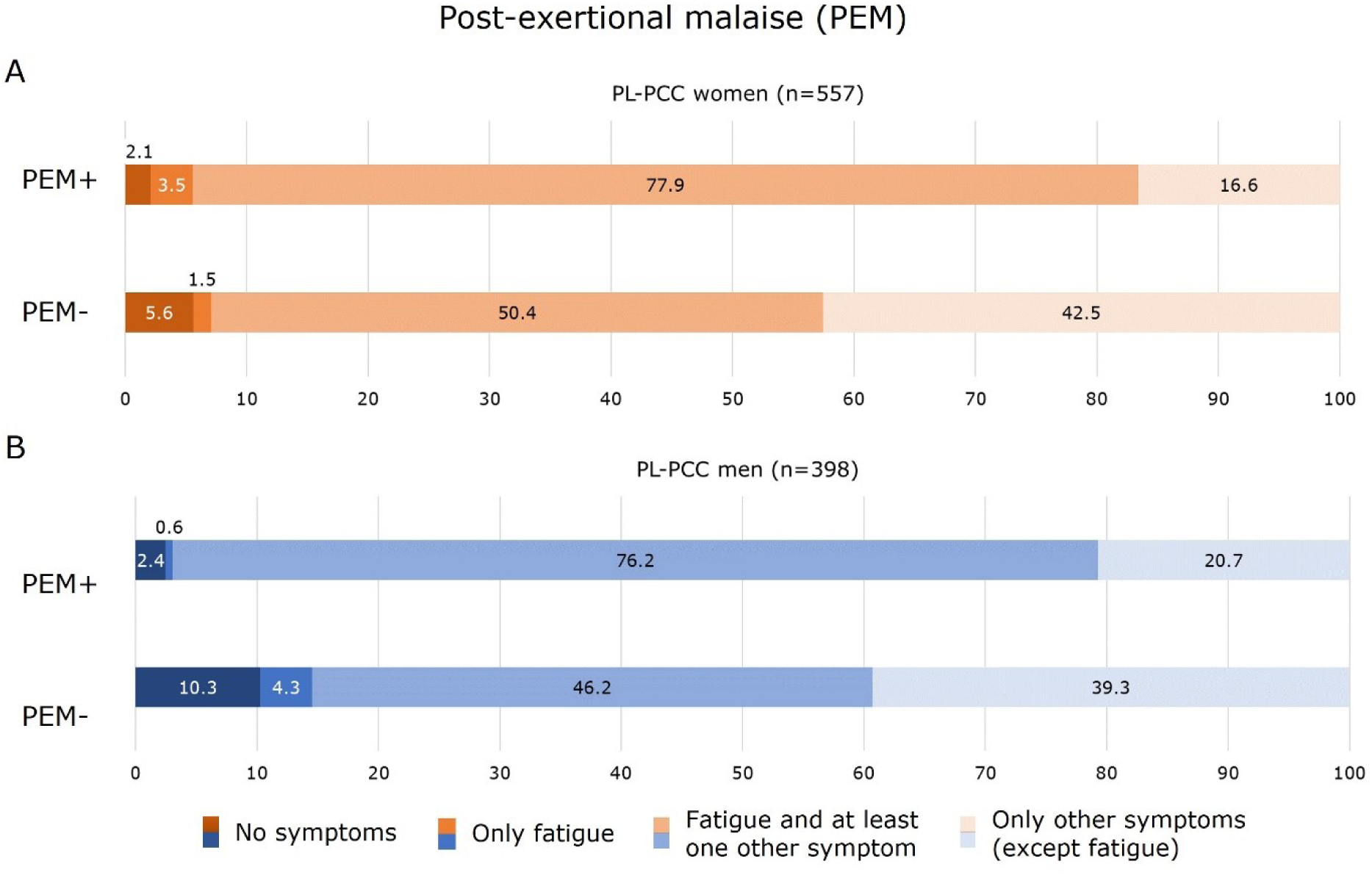
Experienced symptoms for people who were SARS-CoV-2 positive and living with post-COVID-19 condition (PL-PCC) with post-exertional malaise (PEM) and without PEM

Of the PL-PCC women who had OI, 81.0% currently experienced fatigue (of which the majority also experienced other symptoms) (Figure 6A). In PL-PCC men who had OI, 76.6% experienced fatigue and other symptoms (Figure 6B).

**Figure 6.**
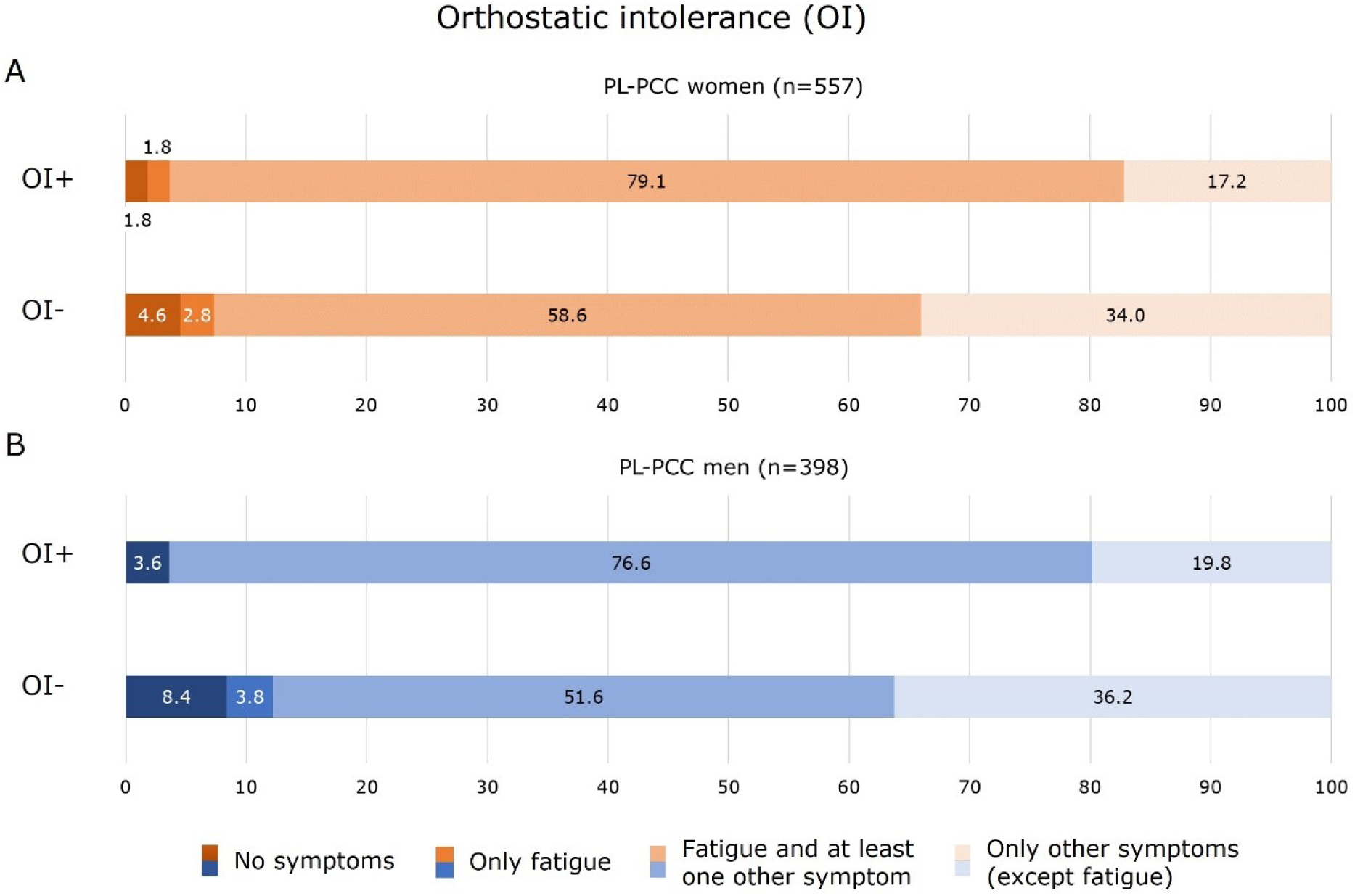
Experiencing symptoms for people who were SARS-CoV-2 positive and living with post-COVID-19 condition (PL-PCC) with orthostatic intolerance (OI) and without OI

## Discussion

Results of the PRIME post-COVID study demonstrate that of people who feel unrecovered since SARS-CoV-2 infection (i.e., defined as PL-PCC), between 48.1% (women) and 41.2% (men) have PEM. Although PEM and OI are identified frequently in both men and women, PEM is more prevalent in PL-PCC women. The proportion of OI is comparable between PL-PCC men (27.9%) and women (29.3%). Of PL-PCC, PEM and OI were co-concurrent in 19.6% and 23.7% of men and women, respectively. Proportions of PEM and OI were notably higher in people of middle or younger age, those who have comorbidities and who live alone. The high proportions of PEM and OI call for standard screening in PL-PCC, regardless whether fatigue is reported, by medical and allied healthcare professionals to avoid inappropriate exercise-based treatment for PL-PCC.

A previous study showed OI estimates to be comparable to the current study with 30.7%, but PEM estimates in PL-PCC to be higher with 81.9% (3). The PEM proportion estimates of the current study are substantially lower (between 41.2% and 48.1%). This might (partly) be explained by different recruitment methods. The current study invited adults being tested and registered in the national COVID-19 registry, thereby recruiting a population-based sample. The previous study partly recruited participants via COVID-19 online support groups, probably including a high proportion of more severe or more aware PCC cases, resulting in selection bias. Besides, a greater proportion of women (78.9%) was included compared to the current study (58.3%), overestimating PEM proportions as PEM is more prevalent in women than men. Furthermore, items used to define PEM differed, as the previous study used only one item. Another study estimated PEM prevalence to be 58.7% in adults experiencing persistent symptoms (≥4 weeks) since infection, using the same validated DSQ-PEM questionnaire (32). They also included a higher share of women (85.5%), however, the prevalence of PEM was more comparable to the results of the current study. As the prevalence of both PEM and OI in the general population is not well known, including a SARS-CoV-2 negative group (as in our study) to estimate background risk is recommended.

The proportions of PEM and OI in negatives were 10.7%-20.4% and 9.9%-10.8%, respectively. Compared to negatives, the PEM and OI proportions were substantially higher in PL-PCC regardless of PCC definition applied (Supplementary Figure 1). Notably, the proportion of PEM was higher in negative women (20.4%) than PnL-PCC women (10.2%; p=0.003). The reason is unknown, however an explanation might be possible misclassification regarding being truly SARS-CoV-2 negative, as we cannot rule out that initially SARS-CoV-2 tested negative people might have been untested and actually be SARS-CoV-2 positive. Besides, the PnL-PCC group explicitly mentioned that they felt recovered since infection, which probably results in a lower chance of having PEM. In the negatives, we did not ask whether they felt recovered after infection, as they did not report a positive SARS-CoV-2 test. Furthermore, we were unable to exclude negatives who already had ME/CFS or fibromyalgia before their SARS-CoV-2 test, which we were able to do for the positives.

The current study revealed that people with comorbidities or those who are living alone had substantially more often PEM or OI, which calls for specific attention to these subgroups when presenting with PCC. Nevertheless, in all subgroups proportion estimates were higher in PL-PCC compared to negatives.

Our results should alert clinicians and allied healthcare professionals to standardly screen for PEM and OI in people who feel unrecovered after SARS-CoV-2 infection or in those who might have PCC. Report of fatigue is not a suitable indicator for PEM or OI to be used in practice (i.e., fatigue is not reported in about one in five PL-PCC who have PEM or OI). Screening should thus be done using appropriate tools. Fortunately, there are simple and easy to use questionnaires available for healthcare professionals to use in daily practice. The DSQ-PEM and DSQ-2 are freely available and composed of only a few questions to validly indicate the presence of PEM or OI. These questionnaires and corresponding cut-off values are currently used in daily practice when diagnosing ME/CFS.

Some strengths and limitations of our study must be discussed. First, the large population-based cohort including SARS-CoV-2 negatives represents the main strength. Until now, studies only included PL-PCC when estimating PEM and OI proportion, without using a SARS-CoV-2 reference group. Second, validated questionnaires were used to assess PEM and OI. Third, we were able to demonstrate that the PEM and OI proportions in PL-PCC were more or less similar when using different PCC definitions, or when examining different periods since infection, stating the robustness of the proportion estimates.

The main limitation is the possibility of selection bias. Only 55.9% of invitees participated in the follow-up questionnaire. It is unknown whether this would lead to over– or underestimation of PEM and OI. Besides, misclassification regarding being truly SARS-CoV-2 negative cannot be ruled out. This could have led to overestimated PEM and OI proportion in the negative group. Furthermore, it is likely that people with most severe PCC, PEM and OI were not included in the questionnaire, since they would be unable to complete the relatively long set of questions. This may cause an underestimation by the current study of the severity and proportion of PEM and OI in people living with PCC.

In conclusion, more attention and better identification of PEM and OI in PL-PCC is urgently needed to tailor treatment strategies, to avoid harmful exercise-based treatments and to promote appropriate and safe therapies. Therefore, we suggest standard screening for PEM and OI, to increase identification by healthcare professionals by using simple questionnaires. Furthermore, special attention should be given to people having comorbidities or living alone.

## Author contributions

DP, MV, CB, SB, KK, CHe, MS, CHo, and ND designed the study and developed the questionnaire. DP wrote the first draft of the manuscript. All authors revised the manuscript critically for important intellectual content, approved the definitive version, and agreed to be accountable for all aspects of the manuscript.

## Acknowledgements

We gratefully acknowledge LCJ Steijvers, CPB Moonen, S Mujakovic, HLG ter Waarbeek, N Bouwmeester-Vincken and AW Vaes for their valuable contribution throughout the shaping and data collection of the PRIME post-COVID study.

## Data availability

Data cannot be shared publicly because the data contains potentially identifying patient information. Data are available on request from the head of the data-archiving South Limburg Public Health Service (contact via) for researchers who meet the criteria for access to confidential data.

