## Supplementary Figure 1 for "High proportions of post-exertional malaise and orthostatic intolerance in people living with post-COVID-19 condition: the PRIME post-COVID study"

Supplementary Figure 1. Post-exertional malaise (PEM) and orthostatic intolerance (OI) proportion estimates according to other, previously used post-COVID-19 condition definitions

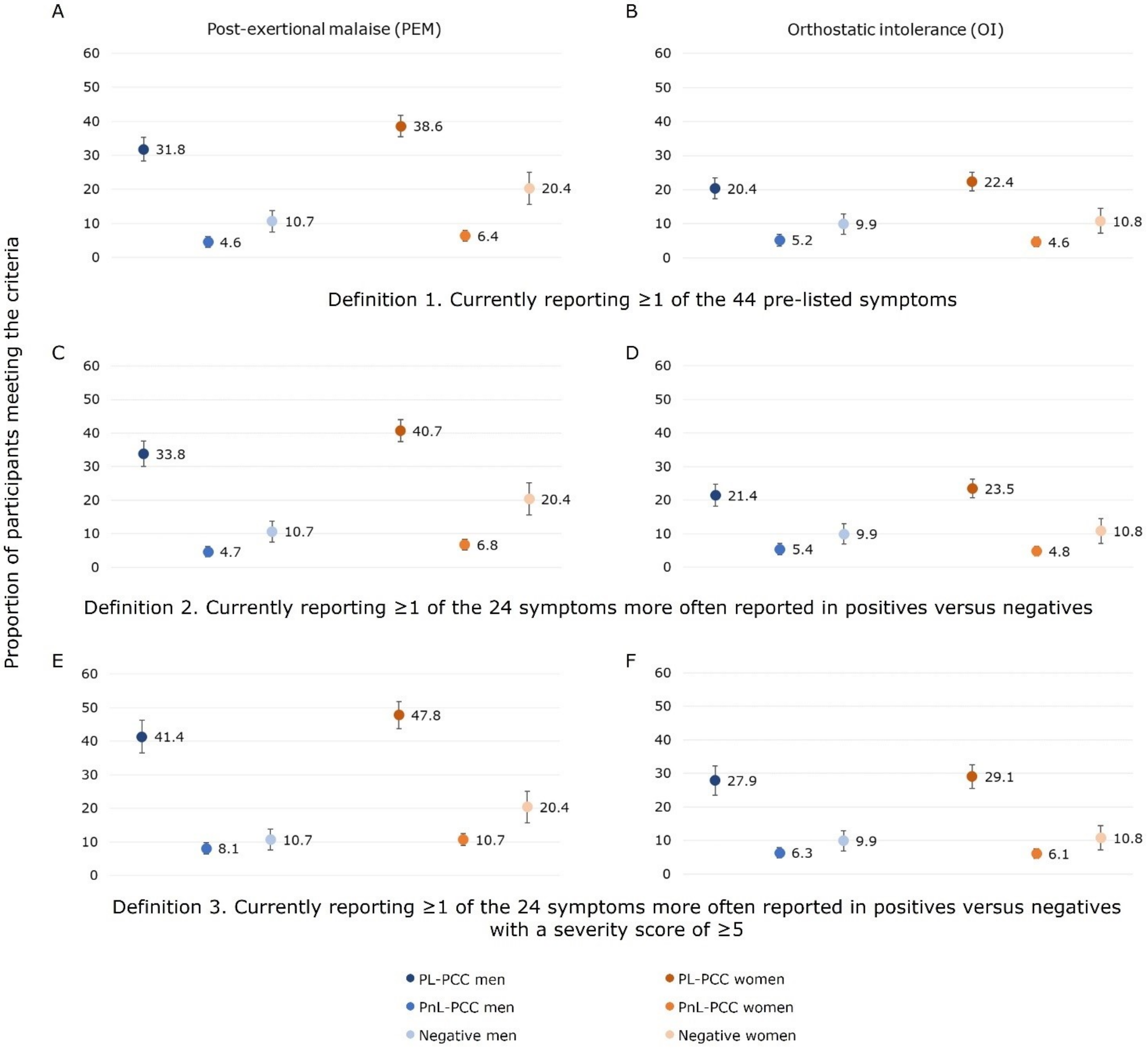
