## Supplementary Table 1 for "High proportions of post-exertional malaise and orthostatic intolerance in people living with post-COVID-19 condition: the PRIME post-COVID study"

Supplementary Table 1. Proportions of post-exertional malaise (PEM) and orthostatic intolerance (OI) in people living with post-COVID-19 condition (PL-PCC) separate for men and women, stratified by various periods since testing

| Months since positive SARS-CoV-2 test | Men (n=398) | | | | | | | Women (n=557) | | | | | | |
|---|---|---|---|---|---|---|---|---|---|---|---|---|---|---|
| | n | % PEM | 95% CI | | % OI | 95% CI | | n | % PEM | 95% CI | | % OI | 95% CI | |
| 3-6 | 25 | 44.0 | 24.5 | 63.5 | 32.0 | 13.7 | 50.3 | 28 | 42.9 | 24.6 | 61.2 | 25.0 | 9.0 | 41.0 |
| 6-9 | 40 | 52.5 | 37.0 | 68.0 | 32.5 | 18.0 | 47.0 | 74 | 56.8 | 45.5 | 68.1 | 35.1 | 24.2 | 46.0 |
| 9-12 | 25 | 40.0 | 20.8 | 59.2 | 24.0 | 7.3 | 40.7 | 46 | 39.1 | 25.0 | 53.2 | 19.6 | 8.1 | 31.1 |
| 12-18 | 182 | 40.1 | 33.0 | 47.2 | 31.3 | 24.6 | 38.0 | 216 | 55.6 | 49.0 | 62.2 | 30.6 | 24.5 | 36.7 |
| >18 | 126 | 38.9 | 30.4 | 47.4 | 21.4 | 14.2 | 28.6 | 193 | 50.3 | 43.2 | 57.4 | 28.5 | 22.1 | 34.9 |

CI: confidence intervals; OI: orthostatic intolerance; PEM: post-exertional malaise.
